# Severe disease is not essential for a high neutralizing antibody response post-SARS-CoV-2 infection

**DOI:** 10.1101/2024.08.21.24312063

**Authors:** Afrah Khairallah, Zesuliwe Jule, Alice Piller, Mallory Bernstein, Kajal Reedoy, Yashica Ganga, Bernadett I. Gosnell, Farina Karim, Yunus Moosa, Thumbi Ndung’u, Khadija Khan, Alex Sigal

## Abstract

Neutralizing antibody responses correlate with protection from SARS-CoV-2 infection, yet higher neutralizing responses associate with more severe disease. Whether people without severe disease can also develop strong neutralizing responses to infection, and the pathways involved, is less clear. We performed a proteomic analysis on sera from 71 individuals infected with ancestral SARS-CoV-2, enrolled during the first South African infection wave. We determined disease severity by whether participants required supplemental oxygen and measured neutralizing antibody levels at convalescence. High neutralizing antibodies were associated with high disease severity, yet 40% of participants with lower disease severity had neutralizing antibody levels comparable to those with severe disease. We found 130 differentially expressed proteins between high and low neutralizers and 40 between people with high versus low disease severity. Five proteins overlapped, including furin, a protease which enhances SARS-CoV-2 infection. High neutralizers with non-severe disease had similar levels of differentially expressed neutralization response proteins to high neutralizers with severe disease, yet similar levels of differentially expressed disease severity proteins to participants with non-severe disease. Furthermore, we could reasonably predict who developed a strong neutralizing response based on a single protein, HSPA8, involved in clathrin pit uncoating. These results indicate that a strong antibody response does not always require severe disease and may involve different pathways.

## Introduction

Many transient viral infections elicit a neutralizing antibody response which not only helps to clear the virus, but also protects convalescent individual from re-infection^1^. This has been shown for SARS-CoV-2, and until the Omicron variant emerged, re-infection was rare^2^. Binding of neutralizing antibodies prevents the viral spike protein from accessing the angiotensin-converting enzyme 2 (ACE2) viral receptor^3^. Neutralizing antibody levels strongly correlate with the degree of vaccine mediated protection^4–7^.

Factors predisposing to higher disease severity and mortality in Covid-19 include male sex, diabetes, hypertension, and HIV^22^. Higher disease severity results in higher neutralizing antibody levels in SARS-CoV-2 infection^8–19^. In contrast, asymptomatic infection associates with a low neutralizing antibody response^20,21^. This opens the question of whether severe disease is necessary for a robust antibody response. An alternative is that both are driven by shared factors such as high viral titers or prolonged infection^23^. Given that severe Covid-19 is due to lower respiratory tract infection, which causes acute respiratory distress syndrome (ARDS) from aberrant inflammation interfering with gas exchange^24,25^, it is possible that a more controlled immune response could avoid ARDS while still being sufficiently robust to elicit high neutralizing antibody levels.

Here, we asked whether severe Covid-19 is required for high neutralizing antibody levels post-infection. To avoid confounding the results with re-infection and vaccination, we selected individuals from our cohort^26^ who were infected by ancestral SARS-CoV-2 in the first Covid-19 infection wave in South Africa, before vaccination was available and variants arose. Given the rarity of reinfection pre-Omicron^27^, the infections we studied were very likely first exposures to SARS-CoV-2. We separated participants with Covid-19 into higher severity versus lower severity by whether participants required supplemental oxygen. The requirement for supplemental oxygen is a key measure in ordinal scales like that used by the World Health Organization^28^. We used proteomics to examine the differentially regulated proteins and pathways in people with different combinations of disease severity and antibody response.

We observed that, while higher severity cases tended to make strong antibody responses, a subset of participants with more mild disease who did not require supplemental oxygen also showed high neutralizing antibody levels. There was minor overlap between the differentially expressed proteins associated with severity versus those associated with the neutralizing antibody response. Additionally, levels of heat shock protein family A member 8 (HSPA8) emerged as predictive of neutralizing responses.

## Results

### Cohort characteristics

We enrolled 72 participants infected during the first SARS-CoV-2 infection wave in Durban South Africa lasting from March to October 2020^29^ (Figure 1, Table 1). One participant in the cohort had immunosuppression due to advanced HIV disease with a CD4 count of 6 at enrollment and an HIV viral load of 34,151 copies/mL. This participant did not make a neutralizing antibody response and had persistent SARS-CoV-2 infection, as described in our previous work^29–31^. Because of the outlier immune state, we excluded this participant from further analysis and analyzed results for 71 participants.

**Figure 1:**
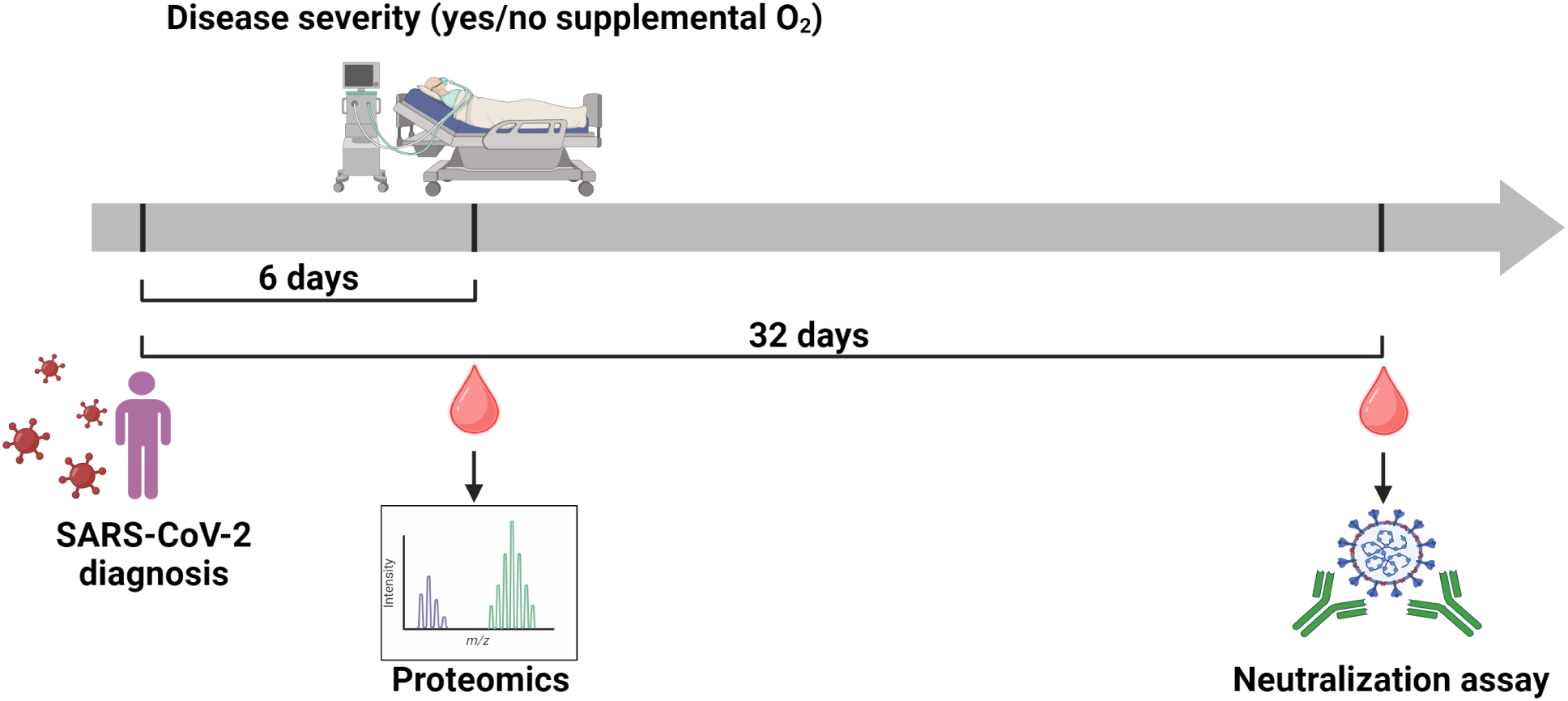
Study design and timeline. Study participants were enrolled in the first South African SARS-CoV-2 infection wave between March and October 2020, where enrollment was a median of 6 days post-diagnosis. At enrollment, disease severity was assessed by whether a participant required supplemental oxygen. During this visit, a blood draw was performed, and blood plasma used for proteomic analysis using SomaScan proteomics. At 28 days post-enrollment, a second blood draw was performed, and blood plasma was used in a live virus neutralization assay against ancestral SARS-CoV-2 to assess neutralization capacity.

**Table 1:** Participant characteristics and comparison of non-severe vs severe groups.

| Parameter | All | Non-severe | Severe | p-value |
| --- | --- | --- | --- | --- |
| n | 72 | 55 | 17 |  |
| Days diag. to enrol., median (IQR) | 6 (4 - 8) | 6 (4 - 8) | 6 (5 - 7) | 0.87 |
| Days diag. to neut. sample, median (IQR) | 32 (17 - 35) | 30 (16 - 35) | 33 (28 - 34) | 0.22 |
| Age in years, median (IQR) | 42 (34 - 50) | 42 (32 - 50) | 40 (36 - 55) | 0.31 |
| People living with HIV, n (%) | 34 (47) | 25 (45) | 9 (53) | 0.78 |
| Sex, male (%) | 22 (31) | 15 (27) | 7 (41) | 0.37 |
| CD4 at enrol., median (IQR) | 590 (340 - 940) | 700 (470 - 1100) | 380 (270 - 590) | 0.0072 |
| CD4 at neut. sample, median (IQR) | 830 (510 - 1100) | 820 (530 - 1100) | 840 (390 - 900) | 0.43 |
| NLR at enrol., median (IQR) | 2 (1.2 - 4.1) | 1.9 (1.2 - 2.5) | 4.2 (1.3 - 7.5) | 0.043 |
| NLR at neut. sample, median (IQR) | 1.6 (1.2 - 2.1) | 1.6 (1.2 - 2.1) | 1.5 (1.2 - 1.9) | 0.77 |
| D614G FRNT<sub>50</sub>, geomean | 230 | 150 | 900 | 0.00052 |

We scored disease severity by whether the participant required supplemental oxygen^26,28^. Participants were classified into the less severe (no supplemental oxygen) and more severe (supplemental oxygen) groups. For brevity, we refer to these groups as “non-severe” and “severe”, although we recognize there may be gradations of severity within each group. Demographics and comorbidities were recorded. We determined HIV status, HIV viral load, and CD4 T cell concentrations in the blood. Diagnosis of SARS-CoV-2 infection was performed by qPCR. Participants were enrolled a median of 6 days (IQR 4-8 days) post-diagnosis. A quantitative SARS-CoV-2 titer was not available. We performed the first sampling at enrollment (Figure 1). This sample was used for proteomic analysis as well as to measure lymphocytes, neutrophils, CD4 T cell numbers, HIV status, and HIV viral load.

A second sample was taken a median of 32 days (IQR 17-35 days) post-diagnosis, at a time when the infection elicited antibody response should be close to its peak^16^. This sample was used to measure neutralizing antibody levels by a live virus focus reduction neutralization test (FRNT^32,33^). Fifty-five participants (76%) were determined to have non-severe disease because they did not require supplemental oxygen, and 17 (24%) were classified as severe. People living with HIV (PLWH) accounted for 47% of the participants, reflecting the high HIV prevalence in the province of KwaZulu-Natal in South Africa where Durban is located^34^.

CD4 T cell count was a median of 590 cells/µL (IQR 340-940) at enrollment over the entire participant group and was significantly lower (p=0.0072 by the Mann-Whitney U test, Table 1) in the severe (380 cells/µL, IQR 270-590) relative to the non-severe (700 cells/µL, IQR 470-1100) group. The lower CD4 count in severe disease is expected because severe disease often results in lymphopenia^35–37^. The CD4 count increased upon convalescence at the second visit to a median of 830 cells/µL (IQR 510-1100), and there was no significant difference between the severe (820 cells/µL, IQR 530-1100) and non-severe (840 cells/µL, IQR 390-900) groups (Table 1).

Increased neutrophils are associated with severe disease^26,37–39^ and the neutrophil to lymphocyte ratio (NLR) was significantly higher in the severe group in the first visit (4.2 vs. 1.9, p=0.04 by the Mann-Whitney U test, see Table 1). The NLR dropped and was similar in both groups at the second, convalescence visit (Table 1). The severe group had significantly higher levels of neutralizing antibodies, as measured by the FRNT_50_, which is the inverse serum dilution in the FRNT assay required to neutralize 50% of ancestral SARS-CoV-2 (Table 1).

### Categorization of participants into high and low neutralizers and association with risk factors

We used the median FRNT_50_ neutralization level (FRNT_50_=358) to categorize participants into high and low neutralizers. Our justification for using the median is that it is simple yet gives two groups which are strongly distinct in neutralization capacity: Geometric mean titer (GMT) FRNT_50_ values were 1348 in the high neutralizer group (n=35) versus 46 in the low neutralizer group (n=36), a 29-fold drop (Figure 2A). The number of participants in the non-severe disease group was 55, out of which 33 were low neutralizers, while 22 were high neutralizers (Table 2). Out of the 16 participants in the high severity group, 13 were high neutralizers while the remaining 3 were low neutralizers (Table 2).

**Figure 2:**
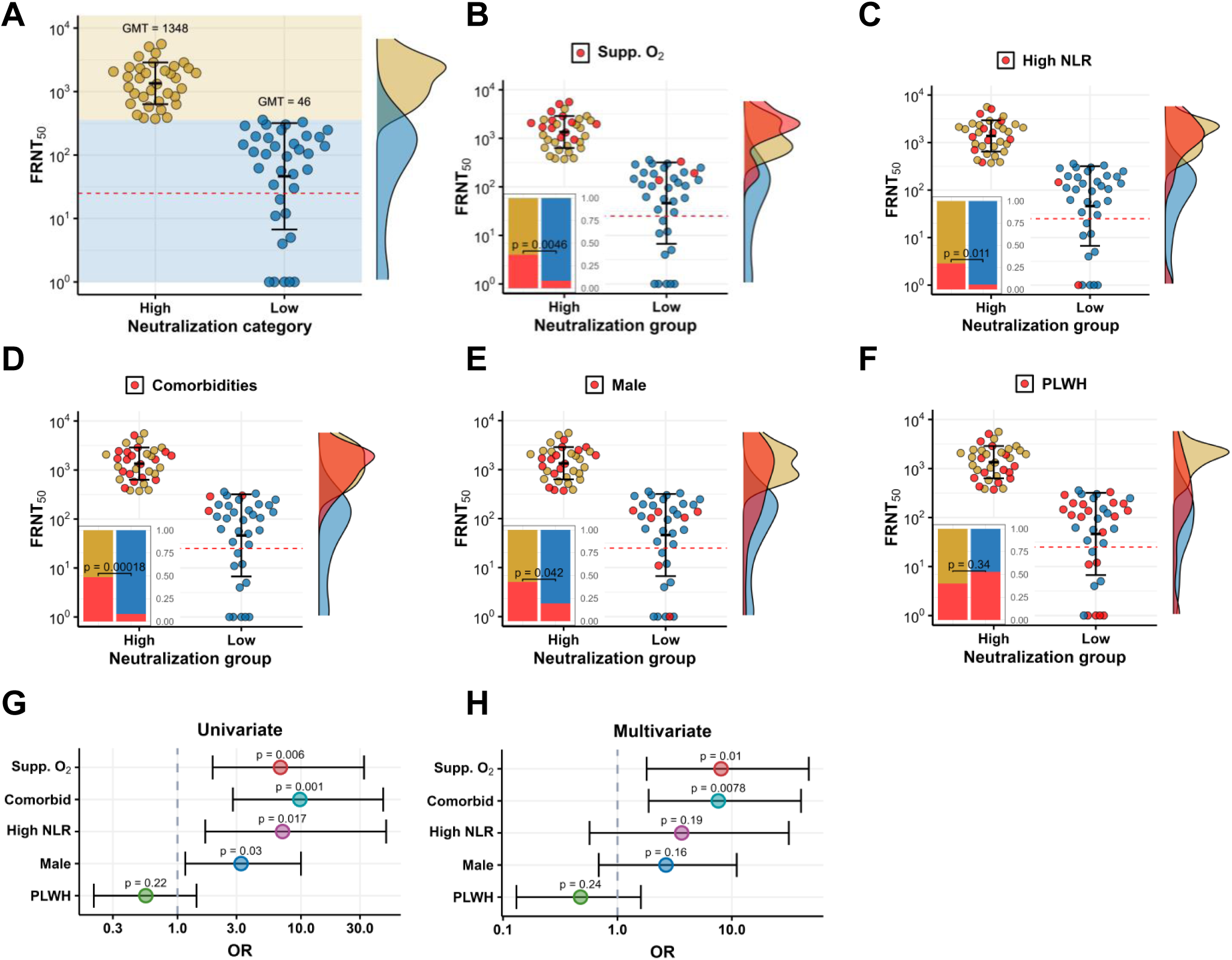
Neutralization capacity associates with disease severity but not HIV status. (A) Participants were divided into high and low neutralizers based on whether the participant’s plasma showed higher or lower neutralizing capacity relative to the median neutralization FRNT_50_ of all participants. For all plots, y-axis shows FRNT_50_, with bar and error bars representing the geometric mean and geometric standard deviation of each group. The horizontal dashed red line marks the limit of quantification (the inverse of the highest plasma concentration used). (B) Participants in the high and low neutralization groups who required supplemental oxygen (red points). Inset shows frequency of participants on supplemental oxygen in each group (p=0.0046 by Fisher’s exact test). (C) High and low neutralizers who had a neutrophil to lymphocyte ratio (NLR) greater than 6. Inset: frequency of high NLR in each group (p=0.011 by Fisher’s exact test). (D) High and low neutralizers who had a comorbidity of diabetes, hypertension, or both. Inset: frequency of comorbidities in each group (p=0.00018 by Fisher’s exact test). (E) High and low neutralizers who were male. Inset: frequency of males in each group (p=0.042 by Fisher’s exact test). (F) High and low neutralizers who were people living with HIV (PLWH). Inset: frequency of PLWH in each group (not significant by Fisher’s exact test). (G) Univariate and (H) multivariate logistic regression odds ratios for odds of having a high neutralization response.

**Table 2:** Participant numbers in neutralization-severity combinations.

|  | Non-severe | Severe | Sum |
| --- | --- | --- | --- |
| Low neutralization | 33 | 3 | 36 |
| High neutralization | 22 | 13 | 35 |
| Sum | 55 | 16 | 71 |

The frequency of participants with high disease severity was significantly higher in the high neutralization group relative to the low neutralization group (p=0.0046 by Fisher’s Exact test, Figure 2B). However, as presented in Table 2, the majority of the 35 participants in the high neutralizer group were non-severe (22 participants, versus 13 with high severity). There was no significant difference in neutralizing antibody levels between participants with high disease severity and high neutralizers with low disease severity (Figure S1).

High NLR, comorbidities, and being male was associated with higher neutralization in univariate analysis. The frequency of individuals with high NLR (>6)^38^, was significantly higher in the high neutralization group (p=0.011 by Fisher’s Exact test, Figure 2C). The frequency of individuals with recorded comorbidities (hypertension and/or diabetes) was also significantly higher in the high neutralizer group (p=0.00018 by Fisher’s Exact test, Figure 2D). Males were significantly more frequent among the high neutralizers (p=0.042 by Fisher’s Exact test, Figure 2E). In contrast, there was no significant difference between the frequency of PLWH versus HIV negative people between the high and low neutralizer groups (Figure 2F). Univariate analysis showed significantly increased odds (odds ratio (OR) >1) of being a high neutralizer with severe disease, comorbidities, high NLR, and being male (Figure 2G). In multivariate analysis, only severe disease and comorbidities remained significant (Figure 2H, with exact numbers and confidence intervals in Table S1). People who both required supplemental oxygen and had comorbidities appeared only in the high neutralizer group (Figure S2). However, the largest subgroup in the high neutralizer group consisted of people with non-severe disease without comorbidities (Figure S2).

### Differentially expressed proteins between high and low neutralizers

We used SomaScan proteomics^40^ to determine the levels of ∼5000 proteins in participant plasma from the blood sample collected at enrollment. Differentially expressed proteins (DEPs) were identified as those with a ≥1.5-fold decrease or increase in mean expression in one group relative to the other, with a p-value corrected by the false discovery rate (FDR) of <0.05^41–43^. We obtained 130 significant DEPs when comparing the high versus the low neutralizer groups (Figure 3A) and 40 DEPs when comparing the non-severe versus the severe disease groups (Figure 3B).

**Figure 3:**
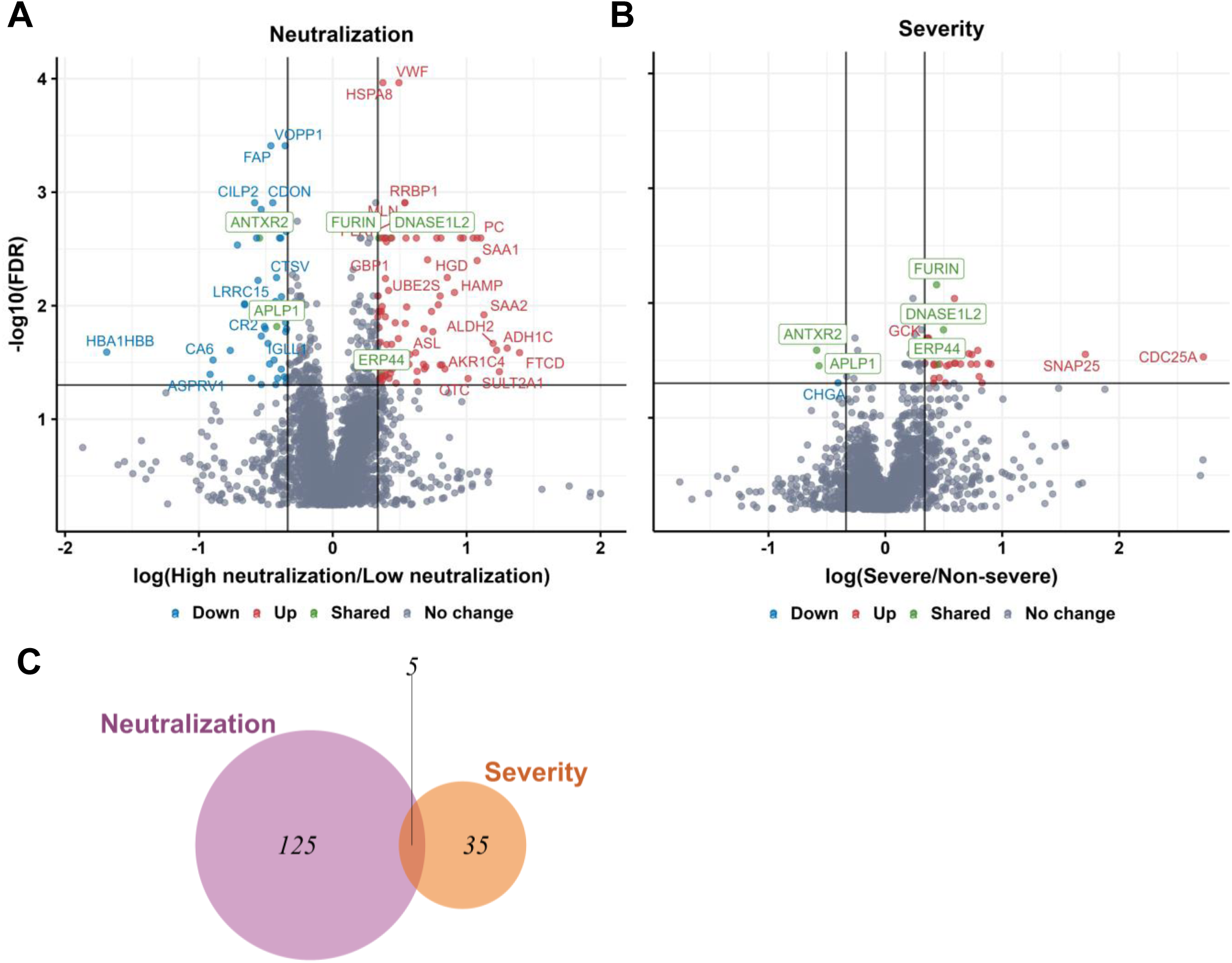
Differentially expressed proteins in neutralization and disease severity. Volcano plots show fold-change versus false discovery rate (FDR) values for each protein in two comparisons: (A) High neutralizers compared to low neutralizers; (B) severe versus non-severe disease. The x-axis represents the log fold-change between the mean protein level values in the group of high versus low neutralizers or high versus low disease severity. Y-axis is the −log_10_ transformed FDR. The vertical lines indicate ±1.5-fold change and the horizontal line FDR=0.05. Significantly differentially expressed proteins are labeled in red (upregulated), blue (downregulated), and green (shared between responses). (C) Venn diagram showing the number of differentially expressed proteins associated with neutralization capacity and disease severity and common to both.

Examples of DEPs with higher levels in high versus low neutralizers (Figure 3A) included HSPA8, also called HSC70^44^. This protein is a chaperone from the HSP70 family of constitutively expressed heat shock proteins and has multiple functions including clathrin uncoating during clathrin-mediated endocytosis^45^, antigen presentation on MHC class II molecules^44,46^, and autophagy^44^. It interacts with proteins from multiple viruses, including the SARS-CoV-2 spike^47^, the influenza M1^48^, and the papillomavirus L2^49^ proteins. Another DEP with significantly higher levels in high neutralizers was von Willebrand factor (VWF), involved in orchestrating the coagulation response^47^.

Furin was an example of a significant DEP in severe versus non-severe disease (Figure 3B). Furin is a protease which promotes SARS-CoV-2 cellular infection by cleaving the S1/S2 polybasic site and therefore facilitating viral fusion^50^. Furin was also elevated in high versus low neutralizers. Another infection promoting factor with higher levels in the high disease severity group was Calpain-2 (CAPN2), involved in positively regulating the cell surface levels of the ACE2 receptor ^51^.

Only five DEPs were found to overlap between the neutralization and disease severity conditions (Figure 3C, proteins marked in green in Figure 3A-B). Upregulated DEPs in common to both high neutralization and high disease severity were furin, DNAse1L2, and endoplasmic reticulum protein 44 (Erp44). Downregulated DEPs in common were anthrax toxin receptor-2 (ANTXR2) and amyloid beta precursor like protein 1 (APLP1). DNAse1L2 has been shown to be transcriptionally activated by inflammatory cytokines^52^ resulting from infection. Erp44, a protein from the thioredoxin family, is upregulated by endoplasmic reticulum stress^53^ and has been reported to be a target of SARS-CoV-2 ORF8^54^. ANTXR2, the receptor for the anthrax toxin, is involved in angiogenesis and cell adhesion^55^. APLP1 is required for glucose homeostasis^56^, and its downregulation is associated with neurological symptoms of Covid-19^57^.

### Gene set enrichment analysis of differentially regulated pathways in neutralization and disease severity

We used Gene Set Enrichment Analysis (GSEA)^58^ to determine the differentially regulated pathways between high and low neutralizers and participants with severe versus non-severe disease (Figure 4). We used the Molecular Signatures Database (MSigDB) Hallmark gene set^59^ and a significance threshold of FDR <0.1 to determine significantly enriched pathways.

**Figure 4.**
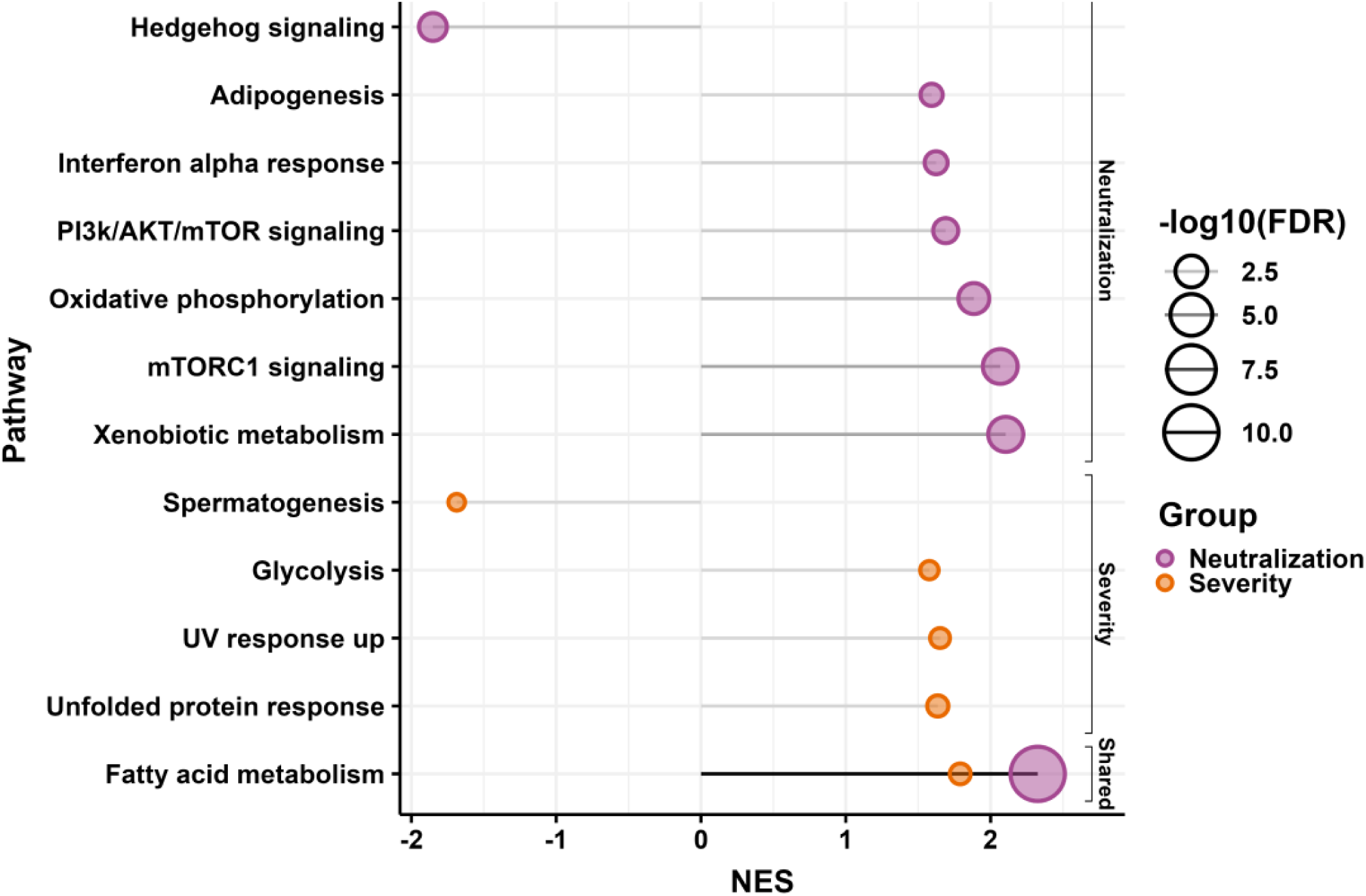
Significantly regulated pathways for neutralization and disease severity. Gene Set Enrichment Analysis (GSEA) results for significantly up or downregulated pathways in participants with high neutralization versus low capacity and those with severe versus non-severe disease. The x-axis represents the Normalized Enrichment Score (NES). Each circle represents a pathway, with the size of the circle corresponding to the -log_10_(FDR) value, with larger circles indicating higher significance. Purple circles represent pathways enriched in the high neutralization group and orange circles represent pathways enriched in the severe disease group. One pathway (fatty acid metabolism) is shared between both groups.

We found 7 upregulated pathways and 1 downregulated pathway in high versus low neutralizers. We found 4 upregulated pathways and 1 downregulated pathway in severe versus non-severe participants. There was one upregulated pathway in common.

In high neutralizers, the upregulated pathways were adipogenesis, the interferon-α (IFN-α) response, PI3k/Akt/mTOR signaling, oxidative phosphorylation, mTORC1 signaling, and xenobiotic metabolism. The downregulated pathway was hedgehog signaling. The pathways upregulated in the high versus low severity groups were glycolysis, the UV response, and the unfolded protein response. Spermatogenesis was downregulated. Similarly to analysis of individual DEPs, there was no extensive overlap in the pathways between responses. The single pathway upregulated in common between the high versus low neutralizers and severe versus non-severe disease was fatty acid metabolism.

### High neutralizers showed similar DEP levels regardless of severity

To better understand differences between high neutralizers with severe versus non-severe disease, we examined the top 20 most significant by FDR DEPs in the neutralization response to determine whether they are similarly or differently regulated in high neutralizers with high versus low disease severity (Figure 5). They included fibroblast activation protein-α (FAP), a serine protease involved in tissue remodeling^60^, motilin (MLN), a hormone promoting gastrointestinal motility^61^, and ribosome-binding protein 1 (RRBP1), involved in the endoplasmic reticulum stress response^62^.

**Figure 5:**
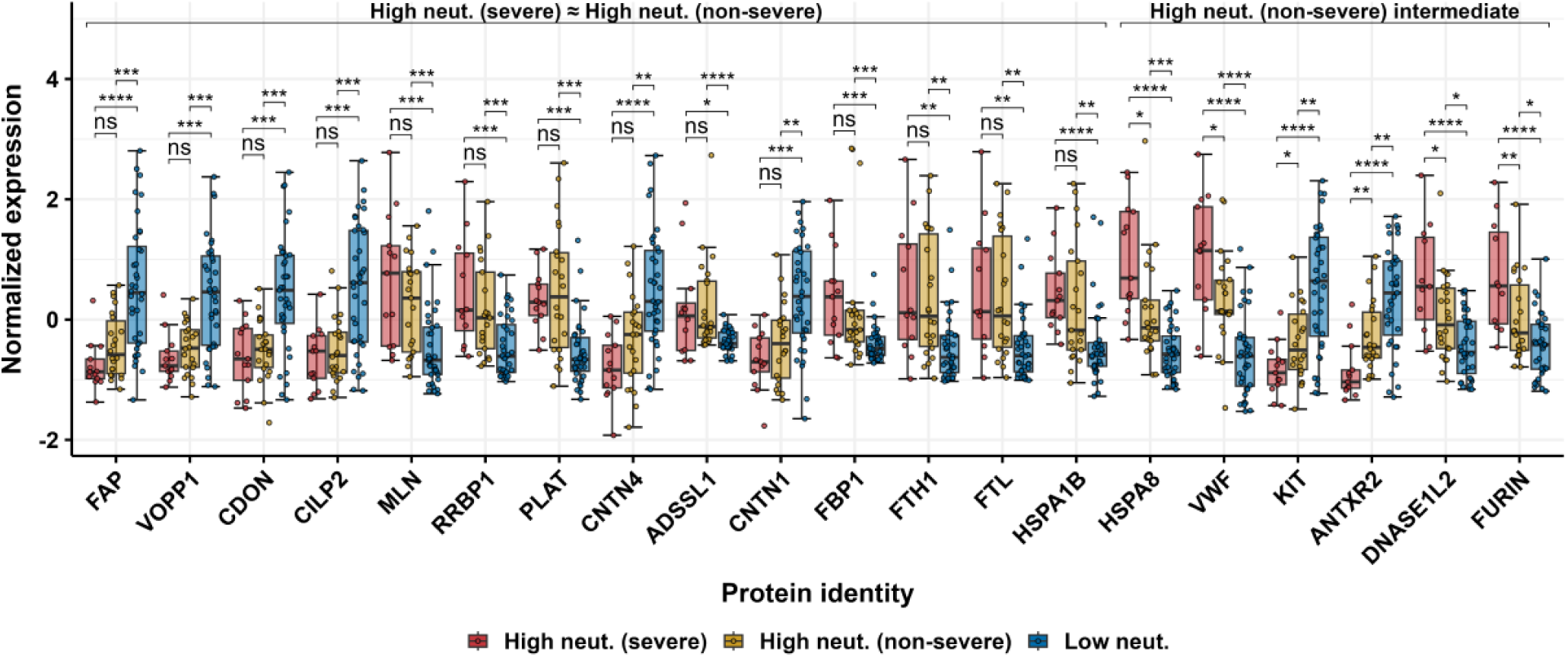
Top differentially expressed proteins in the neutralization response show similar expression in high neutralizers regardless of disease severity. Median Z-score normalized protein levels of the top 20 differentially expressed proteins by FDR are shown for the high neutralizers with severe disease, high neutralizers with non-severe disease, and low neutralizers. Fourteen out of the 20 proteins (left) show no significant difference between the high neutralizers with severe disease versus high neutralizers with non-severe disease. Six proteins (right) show significantly higher levels in high neutralizers with severe disease relative to high neutralizers with non-severe disease. However, the levels of these proteins in low neutralizers are significantly lower still. P-values by the Kruskal-Wallis test with multiple hypothesis correction between group values per protein.

In 14 of these DEPs including FAP, MLN, and RRBP1, levels were not significantly different between high neutralizers with non-severe disease and high neutralizers with severe disease. Both groups had a significantly different level of DEPs relative to the low neutralizer group (Figure 5). For the remaining six proteins, the high neutralizer group with non-severe disease displayed intermediate expression, between high neutralizers with severe disease and the low neutralizers. This group included 3 of the 5 proteins which were in common between the neutralization response and disease severity (furin, ANTXR2, and DNAse1L2), as well as VWF and HSPA8. These results indicate that high neutralizers have levels of the 20 most significant neutralization related DEPs which are generally similar regardless of disease severity.

### HSPA8 level predicts whether infection will elicit high levels of neutralizing antibodies

We investigated whether we could predict strong neutralizing antibody responses based on the DEPs significantly associated with neutralization level. Participants were split into training (60%, n=42) and test (40%, n=29) groups. Significantly regulated proteins were determined by the same FDR and fold-change cut-off as used in the full set analysis (FDR<0.05, fold change≥1.5), resulting in 12 significantly regulated proteins in the neutralization response in the training set (Figure S3). Repeated stepwise regression using bootstrapping was performed to rank the predictive power of each of the 12 proteins by iteratively subtracting or adding each of the proteins from/to the model. Stepwise regression was performed until the Akaike information criterion (AIC), a measure of the trade-off between goodness-of-fit and model complexity was optimized. The top three predictive proteins in order of significance were HSPA8, MLN, and FAP. They were combined in a multivariate logistic regression model (Figure 6A) and analyzed singly in univariate regression (Figure 6B-D).

**Figure 6:**
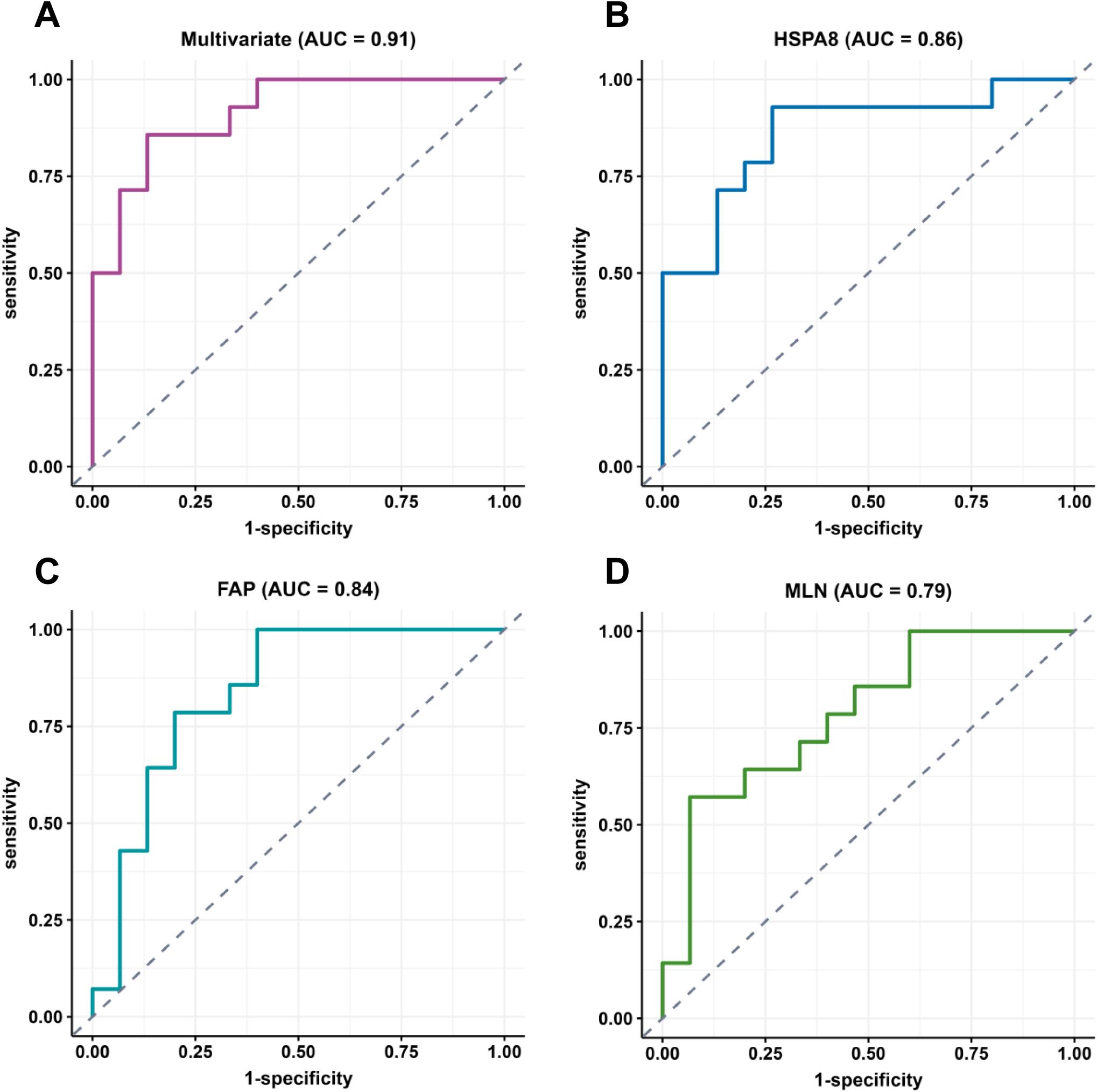
Predictive models classifying participants into high versus low neutralization groups. Shown are ROC curves for (A) multivariate (HSPA8+FAP+MLN), (B) HSPA8, (C) FAP, and (D) MLN. Areas Under the Curve (AUC) were 0.91 (multivariate), 0.86 (HSP8A), 0.84 (FAP), and 0.79 (MLN). Dashed diagonal line represents the performance of a random classifier with AUC=0.50.

The combination of HSPA8, MLN, and FAP resulted in good discrimination between high neutralizers versus low neutralizers, showing an area under the curve (AUC) of 0.91 (Figure 6A). Using HSPA8 alone, the model could reasonably distinguish between low and high neutralization outcomes in the test group (AUC=0.86, p=0.0018, Figure 6B). The predictive power of FAP (AUC=0.84) and MLN (AUC=0.79) was slightly lower (Figure 6C-D).

### Similar severity related protein levels in high and low neutralizers with non-severe disease

Lastly, to test whether the similarity in disease severity between low and high neutralizers with non-severe disease was supported by the similarity in protein expression of severity-related proteins, we examined the top 20 significant by FDR proteins that were differentially expressed in the severe versus the non-severe groups. Included in this group was CAPN2, which increases the levels of the SARS-CoV-2 receptor ACE2^51^, and CD79A, a marker of B cell activation reported to be upregulated upon SARS-CoV-2 infection^63^.

We found that for 16 out of the top 20 DEPs examined, the expression level was not significantly different between the high and low neutralizers among the participants with non-severe disease (Figure 7). In addition, in all but two of the 16 proteins, there was a significant difference between both the high and low neutralizers with non-severe disease and the severe disease group. In 3 out of the remaining 4 DEPs which did not follow this pattern (furin, ANTXR2, and DNAse1L2), the non-severe high neutralizers had intermediate protein levels which fell between the non-severe low neutralizers and the severe disease group. Interestingly, these DEPs were also common to both the severity and neutralization responses (Figure 3A-B). The fourth DEP, olfactomedin 2 (OLFM2), a protein mostly expressed in neurons and found to regulate metabolism^64^, did not show a significant difference between the non-severe high neutralizers and the severe disease group (Figure 7). These results indicate that, for the majority of the top 20 DEPs by significance which distinguished severe from non-severe disease participants, levels were similar between non-severe participants, regardless of participant neutralization capacity.

**Figure 7:**
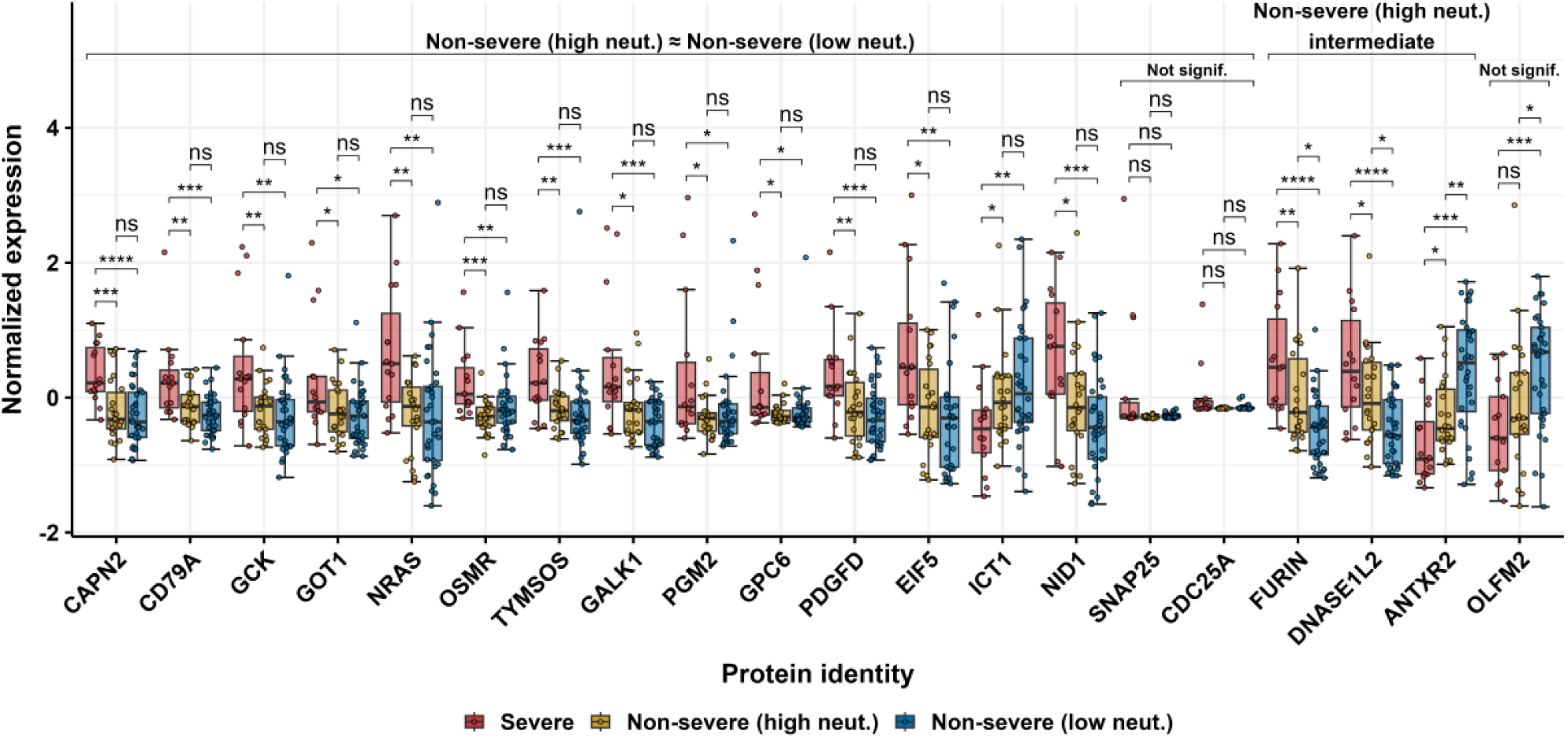
Top differentially expressed proteins in the disease severity response show similar expression regardless of neutralization capacity. Median Z-score normalized protein of the top 20 differentially expressed proteins by FDR are shown for the high severity group and the low severity group, with the latter divided into high and low neutralizers. Sixteen out of the 20 proteins (left) show no significant difference between non-severe high versus low neutralizers. Three proteins (middle-right) show levels in the low severity, high neutralizer group which are intermediate between participants with high severity and low severity and low neutralization. Levels of the remaining one protein show no significant difference between participants with severe disease and high neutralizers with non-severe disease. p-values by the Kruskal-Wallis test with multiple hypothesis correction between group values per protein.

## Discussion

We investigated disease severity, proteomic profiles, and infection-elicited neutralizing antibody levels in a cohort of 71 South African people infected with SARS-CoV-2 during the first ancestral SARS-CoV-2 infection wave. We stratified our cohort into severe and non-severe disease based on whether participants required supplemental oxygen and separated participants into high neutralizers and low neutralizers at convalescence based on their plasma neutralization capacity of ancestral SARS-CoV-2. We found that high disease severity and comorbidities were significantly associated with high neutralization capacity. However, there were participants with non-severe disease who had had similarly high neutralizing antibody levels to those with severe disease. These non-severe high neutralizers showed similar differentially expressed neutralization response protein levels to high neutralizers with severe disease. Meanwhile, their levels of differentially expressed proteins involved in disease severity were similar to participants with non-severe disease.

The enriched pathways we observed in high versus low neutralizers were generally associated with viral replication or the immune response. These included the interferon-α (IFN-α) response, a key part of the innate immune response to viral infections, with rapid production of IFN-α triggering antiviral and pro-inflammatory effects^65^; PI3k/Akt/mTOR signaling, which is is a mediator of cell cycle progression and cell survival^66^, with activation increasing viral replication^67–69^; Oxidative phosphorylation (OXPHOS), the process by which mitochondria generate ATP, with SARS-CoV-2 infection has been reported to promote OXPHOS and increases ATP production^70^; the mTORC1 pathway, a metabolic regulator of cell growth^71^, with mTORC1 inhibitors shown to reduce SARS-CoV-2 replication^72^; hedgehog signaling, involved in development, cell proliferation, survival, and immune regulation, and modulated by multiple viruses^73^; fatty acid metabolism, the pathway in common between the neutralization response and disease severity, essential for the replication of enveloped viruses^74–76^. The other enriched pathways in the neutralization response may be associated with higher disease severity, although they do not come up as enriched pathways in the severe versus non-severe disease analysis: adipogenesis, associated with obesity, a known risk factor for severe Covid-19^77–80^, and upregulation of xenobiotic metabolism, which may indicate the presence of pharmacological interventions^81^.

Like with the neutralization response, the enriched pathways in severe versus non-severe disease show links to SARS-CoV-2 replication. However, some make sense as consequences of severe disease independently of such replication. Thus, the upregulated pathway of glycolysis increases the replication of SARS-CoV-2 and other viruses^82–84^. However, hypoxia, which may have been present in participants with respiratory distress who required supplemental oxygen, also results in a metabolic switch from mitochondrial respiration to increased glycolysis^85^. The unfolded protein response may be upregulated because of the increased production of improperly folded proteins due to ER stress during SARS-CoV-2 cellular infection^86^. Interestingly, ER stress and misfolded proteins have also been linked to autoimmune disease^87^, which has been reported to be elevated post-Covid-19^88^. Upregulation of proteins involved in the UV response may be because SARS-CoV-2 induces cell cycle arrest to gain cellular resources^89^. Decreased spermatogenesis, on the other hand, has been previously shown to be a consequence of severe Covid-19 leading to death^91^.

For the neutralization response, differentially expressed proteins were measured at an earlier point in time than the neutralizing antibody response is thought to develop^92^. Therefore, they may be predictive. We found that three proteins, HSPA8, MLN, and FAP, could predict who will be high neutralizers. The best single protein predictor was HSPA8. This protein has multiple roles in cellular homeostasis, including a key role in the uncoating of endocytosed clathrin vesicles^93^. Interestingly, clathrin mediated endocytosis is a major cellular infection pathway for SARS-CoV-2^94^ and facilitation of this process by HSPA8 may increase viral replication. In addition, HSPA8 has an important role in antigen presentation on MHC class II molecules^44,46^, a necessary step in the CD4 helper T cell - B cell interactions which are responsible for the production of effective neutralizing antibodies^95^.

There was minor overlap between differentially expressed proteins and pathways involved in the neutralization response and the disease severity response. The minor overlap argues against the direct dependence of neutralizing antibody levels on disease severity and supports the proposition that a common factor such as high or prolonged viral replication results in both higher infection-elicited neutralization capacity and higher disease severity. However, a limitation to this conclusion is that, because of small group sizes, some differentially expressed proteins did not reach statistical significance in the disease severity response. For example, coagulation factor VWF was significantly higher in high neutralizers (FDR = 0.0001) but narrowly missed statistical significance in disease severity (FDR = 0.055). VWF was previously reported to be upregulated in high disease severity^96^. A second limitation is that our single measure, the requirement for supplemental oxygen, does not capture the range of more versus less severe disease, and therefore disease severity for some participants is misclassified. Nevertheless, while supplemental oxygen is not a perfect measure, most people with severe disease have respiratory failure, although they may die from multiorgan failure or other reasons^97,98^. The presence of non-severe high neutralizers, who have similar neutralization response related protein profiles to severe high neutralizers, may mean that high disease severity is not essential to elicit a robust neutralizing antibody response.

## Methods

### Informed consent and ethical statement

This was an observational study with longitudinal sample collection. The nasopharyngeal swab used to isolate ancestral SARS-CoV-2 as well as all blood samples were obtained after written informed consent from adults with PCR-confirmed SARS-CoV-2 infection enrolled in a prospective cohort of SARS-CoV-2 infected individuals at the Africa Health Research Institute. The study protocol was approved by the Biomedical Research Ethics Committee at the University of KwaZulu-Natal (reference BREC/00001275/2020). Participants were reimbursed for each visit based on time, inconvenience and expenses as approved in the protocol.

### Clinical laboratory testing

CD4 T cell count and HIV viral load quantification were performed from a 4mL EDTA tube of blood at an accredited diagnostic laboratory (Ampath for CD4 and Molecular Diagnostic Services for HIV viral load, both based in Durban, South Africa).

### Cells

The VeroE6 cells expressing TMPRSS2 and ACE2 (Vero E6-TMPRSS2), originally BEI Resources, NR-54970, were used for virus expansion and live virus neutralization assays. The cell line was propagated in growth medium consisting of Dulbecco’s Modified Eagle Medium (DMEM, Gibco 41965-039) with 10% fetal bovine serum (Hyclone, SV30160.03) containing 10mM of hydroxyethylpiperazine ethanesulfonic acid (HEPES, Lonza, 17-737E), 1mM sodium pyruvate (Gibco, 11360-039), 2mM L-glutamine (Lonza BE17-605E) and 0.1mM nonessential amino acids (Lonza 13-114E).

### Live virus neutralization assay (focus reduction neutralization assay)

For all neutralization assays, viral input was 100 focus forming units per well of a 96-well plate. VeroE6-TMPRSS2 cells were plated in a 96-well plate (Corning) at 30,000 cells per well 1 day pre-infection. Plasma was separated from EDTA-anticoagulated blood by centrifugation at 500 × *g* for 10 min and stored at −80 °C. Aliquots of plasma samples were heat-inactivated at 56 °C for 30 minutes and clarified by centrifugation at 10,000 × g for 5 minutes. Virus stocks were added to serially diluted plasma in a 96-well plate (Corning) and antibody–virus mixtures were incubated for 1 h at 37 °C, 5% CO_2_. Cells were infected with 100 μL of the virus–antibody mixtures for 1 h, then 100 μL of a 1X RPMI 1640 (Sigma-Aldrich, R6504), 1.5% carboxymethylcellulose (Sigma-Aldrich, C4888) overlay was added without removing the inoculum. Cells were fixed 20 h post-infection using 4% PFA (Sigma-Aldrich, P6148) for 20 min. Foci were stained with a rabbit anti-spike monoclonal antibody (BS-R2B12, GenScript A02058) at 0.5 μg/mL in a permeabilization buffer containing 0.1% saponin (Sigma-Aldrich, S7900), 0.1% BSA (Biowest, P6154) and 0.05% Tween-20 (Sigma-Aldrich, P9416) in PBS for 2 h at room temperature with shaking, then washed with wash buffer containing 0.05% Tween-20 in PBS. Secondary goat anti-rabbit HRP conjugated antibody (Abcam ab205718) was added at 1 μg/mL and incubated for 2 h at room temperature with shaking. TrueBlue peroxidase substrate (SeraCare 5510-0030) was then added at 50 μL per well and incubated for 20 min at room temperature. Plates were imaged in an ImmunoSpot Ultra-V S6-02-6140 Analyzer ELISPOT instrument with BioSpot Professional built-in image analysis (C.T.L).

### Statistics and fitting

All statistics were performed in GraphPad Prism version 9.4.1. All fitting to determine FRNT_50_ and linear regression was performed using custom code in MATLAB v.2019b (FRNT_50_) or the fit lm function for linear regression, which was also used to determine goodness-of-fit (R^2^) as well as p-value by F-test of the linear model. Limits of quantification were between 1:25 (most concentrated plasma used and 1:3200 (most dilute plasma used)

Neutralization data were fit to:

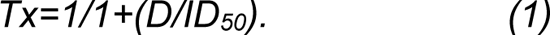

Here Tx is the number of foci at plasma dilution D normalized to the number of foci in the absence of plasma on the same plate. ID_50_ is the plasma dilution, giving 50% neutralization. FRNT_50_ = 1/ID_50_. Values of FRNT_50_ <1 are set to 1 (undiluted), the lowest measurable value. We note that FRNT_50_ < 25 or FRNT_50_ > 3200 fell outside of the dilution series used and were extrapolated from the fit.

### Plasma proteomic profiling

Proteomic analysis of the plasma samples was performed by SomaLogic, Inc. (Boulder, CO, USA) using the SomaScan v4.0 platform^99^. The SomaScan measures were reported as relative fluorescence units (RFU) in a summary ADAT file. These data were then merged with our metadata. Quantile normalization and log transformation were performed on all RFU-reported data to ensure comparability and normalization across samples.

### Differential protein analysis

The proteome changes attributable to both neutralization capacity and disease severity were derived from comparisons between individuals with high versus low neutralization capacity and severe versus non-severe outcomes, respectively. Protein data were log-transformed before testing the difference in means between comparison groups using the Student’s t-test. P-values were corrected to control for the false discovery rate (FDR)^100^. The absolute fold change was rounded up to one decimal place (i.e, fold change values between 1.4 and 1.5 were rounded to 1.5). Proteins with an adjusted p-value < 0.05 and absolute fold change ≥1.5 were considered differentially expressed. These differentially expressed proteins were then visualized using a volcano plot in the R Project for statistical computing and graphical representation.

### Gene set enrichment analysis

Gene Set Enrichment Analysis (GSEA) was performed using the Broad Institute GSEA software version 4.3.3^58,101^, the MSigDB Hallmark gene sets (v2023.2), and the UniProt Human Collection chip platform. The GSEA software was downloaded from https://www.gsea-msigdb.org/gsea/downloads.jsp, and GSEA was performed using default parameter settings except for Number of permutations = 10,000; Permutation type = gene_set; 10 ≤ gene set size ≤ 500.

### Analysis of top variable proteins in the neutralization and severity responses

The top 20 variable proteins from the differential protein analysis of the neutralization response were selected based on FDR value. The proteins were standardized using Z-score normalization, where the mean level of a given protein is subtracted from each value of that protein, and the difference is divided by the protein’s standard deviation. Pairwise differences in median protein levels between high neutralizers with severe disease, high neutralizers with non-severe disease, and low neutralizers (severe and non-severe) were assessed using Mann-Whitney U tests, with p-values adjusted for multiple comparisons using the Benjamini-Hochberg method. Adjusted p-values < 0.05 were marked as significant. Outliers (absolute Z-score > 3) were included in the analysis but excluded from the plot for clarity of the visualization. The same analysis was carried out on the top 20 variable proteins in the severity response, except pairwise differences assessed were between severe disease (high and low neutralizers), non-severe high neutralizers, and non-severe low neutralizers.

### Logistic regression prediction model of neutralization outcome

Participants were split into training (60%, n=42) and testing (40%, n=29) sets. To identify a subset of proteins with the most potential predictive power of neutralization outcome, the training data were subjected to differential protein analysis as described above. This yielded 12 significantly differentially expressed proteins (DEPs) between high and low neutralizers in the training data. The protein data in the training and testing sets were log-transformed and scaled by subtracting the mean and dividing by the standard deviation, with the mean and standard deviation of the training data used for both the training and testing sets. Using bootstrapping (1000 iterations with 100 participant values drawn with replacement per iteration), the neutralization response was iteratively modeled against the 12 DEPs and subjected to forward and backward stepwise regression, selecting the most significant predictors in a regression model by adding (forward) or subtracting (backward) predictors in a stepwise manner, optimizing for Akaike’s information criterion (AIC). With each bootstrap iteration, the proteins selected by stepwise regression were added to a running tally, which served to rank the predictive power of the 12 DEPs upon completion of the bootstrapping. Neutralization response was fitted against the top three ranked proteins, MLN, FAP, and HSPA8, in a binomial logistic regression model, which was trained using the training data. The performance of this multivariate model and the constituent univariate models was assessed using the testing data of ROC curves, and AUC statistics were generated using the R pROC package.

## Author contributions

A.S., A.K., and K.K. conceived the study and designed the study and experiments. Z.J., K.R. Y.G., B.I.G, F.K., Y.M., and K.K identified and provided samples and performed the neutralization experiments. T.N. provided resources. A.P., A.K., and A.S., and M.B. analyzed the data. A.S., A.P., and A.K. prepared the manuscript with input from all authors.

## Data Availability

Sequence of isolated SARS-CoV-2 used in this study has been deposited in GISAID and GenBank with accession numbers EPI_ISL_602626.1 (GISAID), OP090658 (GenBank). It is available upon reasonable request. All R-scripts used in the analysis have been deposited to GitHub (https://github.com/Afrah-Khairallah/Omics-).

## Acknowledgements

We thank Clare Paterson and the team at SomaLogic for performing the proteomics and for helpful comments on the manuscript.

---

**Figure S1:**
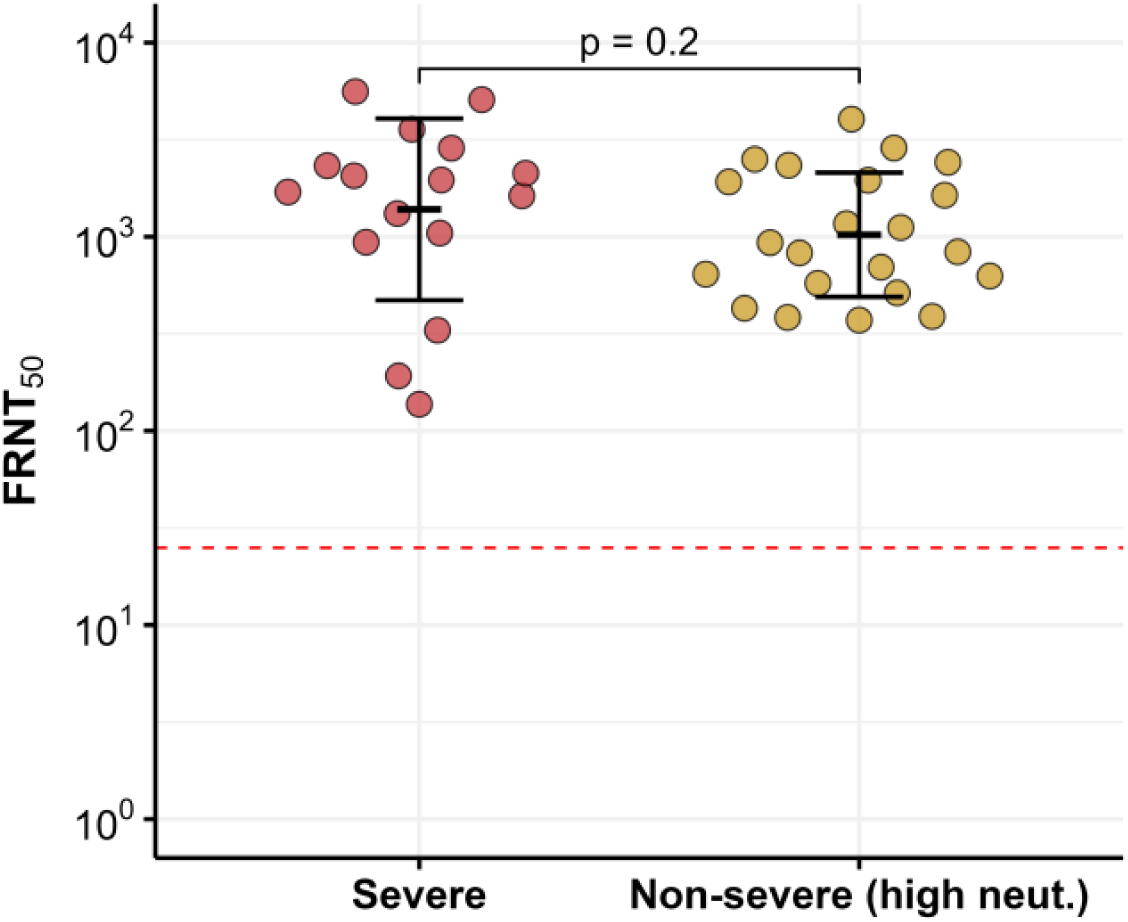
Comparison of neutralization capacity of high neutralizers with low severity to participants with high disease severity. The y-axis shows FRNT_50_, with bar and error bars representing the geometric mean and geometric standard deviation of each group. The horizontal dashed red line marks the limit of quantification (the inverse of the highest plasma concentration used). p-value by the non-parametric Mann-Whitney test.

**Figure S2:**
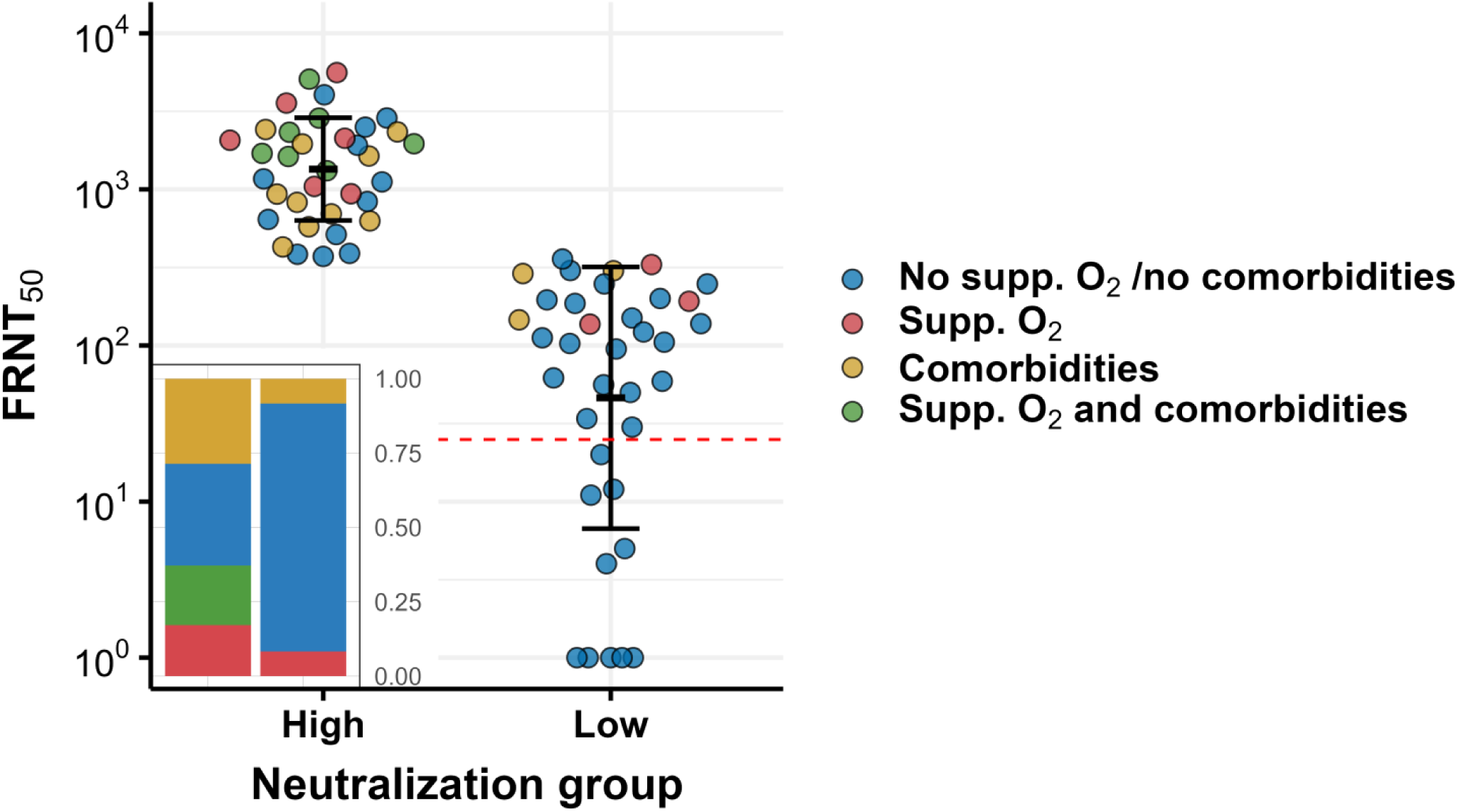
Overlap of comorbidities and requirement for supplemental oxygen in high versus low neutralizers. Participants not requiring supplemental oxygen and having neither diabetes nor hypertension are marked in blue, those requiring supplemental oxygen but without diabetes or hypertension are in red, participants with diabetes or hypertension comorbidities only are in yellow, and those requiring both supplemental oxygen and with diabetes or hypertension comorbidities are in green. Inset shows relative frequencies of the four groups in high versus low neutralizers.

**Figure S3:**
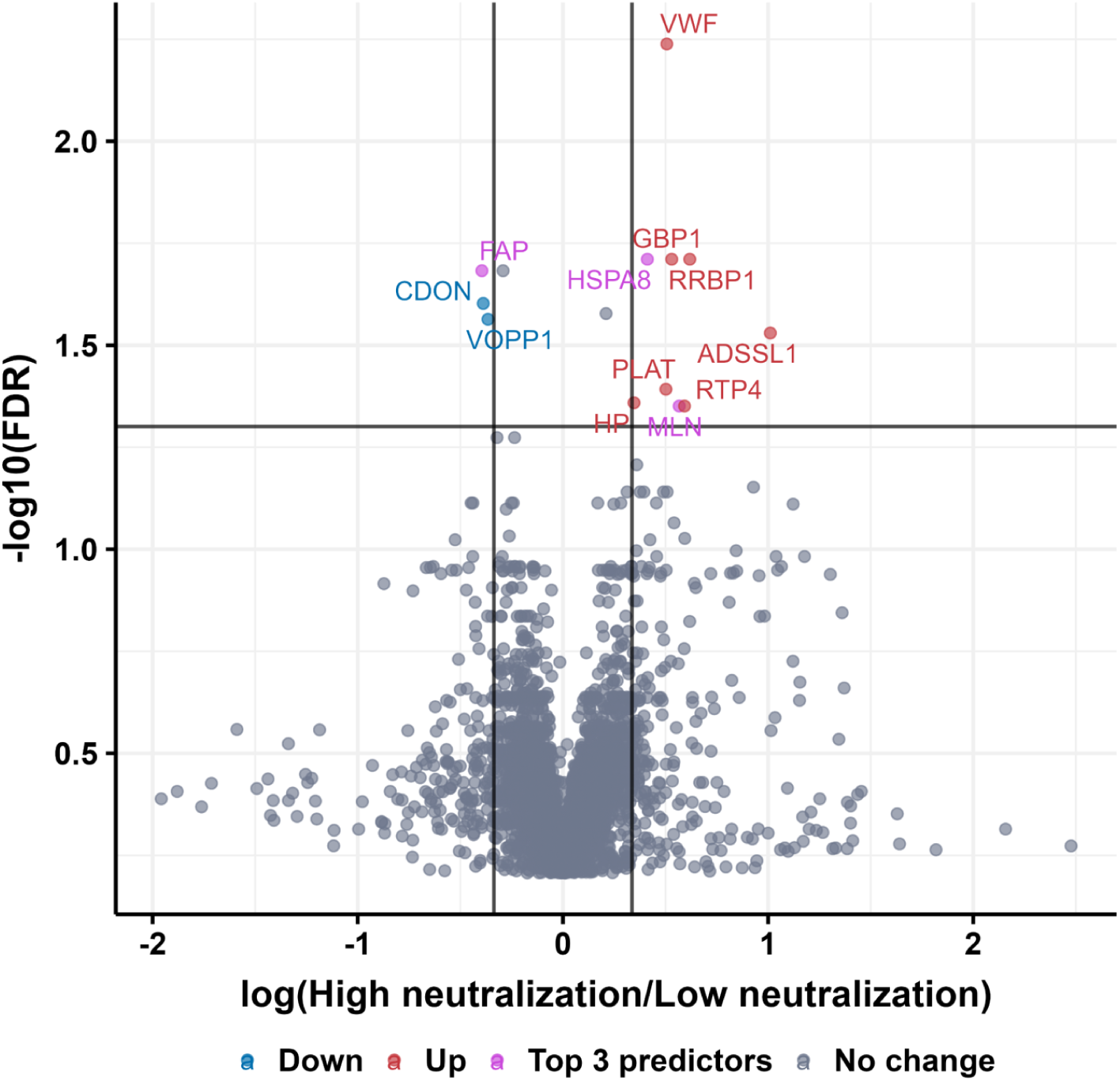
Significantly differentially expressed proteins in the training set. Volcano plots show fold change versus false discovery rate (FDR) values for each protein. The x-axis represents the log fold-change between the mean protein level values in the group of high versus low neutralizers or high versus low disease severity. Y-axis is the −log_10_ transformed FDR. The vertical lines indicate ±1.5-fold change and the horizontal line FDR=0.05. Significantly differentially expressed proteins are labeled in red (upregulated) or blue (downregulated). The proteins showing highest predictive value in the model are highlighted in purple.

**Table S1:** Logistic regression analysis results.

|  | Univariate |  | Multivariate |  |
| --- | --- | --- | --- | --- |
|  | OR (95% CI) | p-value | OR (95% CI) | p-value |
| Supplemental O<sub>2</sub> | 6.81 (1.92; 32.3) | 0.00604 | 8.04 (1.8; 47) | 0.0105 |
| Comorbidities | 9.78 (2.81; 46.2) | 0.00102 | 7.61 (1.87; 40.2) | 0.00781 |
| High NLR | 7.08 (1.68; 48.9) | 0.0169 | 3.64 (0.571; 31.5) | 0.19 |
| Male | 3.27 (1.16; 9.99) | 0.0296 | 2.66 (0.686; 11) | 0.162 |
| PLWH | 0.554 (0.21; 1.43) | 0.224 | 0.476 (0.131; 1.6) | 0.238 |

